# VASCULAR AGE CALCULATION FROM SCORE2 AND SCORE2-OP CARDIOVASCULAR RISK TABLES

**DOI:** 10.1101/2025.05.05.25327022

**Authors:** Jose I. Cuende, Carlos Guijarro

## Abstract

**Aims:** SCORE2 and SCORE2(OP) cardiovascular risk tables were published in 2021 and incorporated into european cardiovascular prevention guidelines, replacing the previous SCORE tables. Since the 2012 version of these guidelines, the concept of vascular age has been included. Although there are vascular age tables derived from SCORE, there are no vascular age tables derived from SCORE2 and SCORE2(OP). The aim of this study is to construct new vascular age tables derived from SCORE2 and SCORE2(OP).

**Methods:** Based on the data from the SCORE2 and SCORE2(OP) risk tables, 8 regressions between age and risk are calculated in baseline conditions for the 8 combinations of sex and cardiovascular risk zones. Another 248 regressions corresponding to the different non-baseline situations are calculated. By equating the risk in each box of the risk tables with the same risk in baseline conditions, the vascular age in each box of the risk tables can be obtained. Regression, determination and intraclass correlation coefficients are calculated to asses adjustment and agreement.

**Results:** Four vascular age tables have been obtained corresponding to countries with low, moderate, high and very high cardiovascular risk. The concordance in vascular age between the 4 tables is greater than 0.99, thus a universal vascular age table has been constructed. Vascular age does not need calibration unlike absolute risk.

**Conclusions:** Vascular age tables have been created from the SCORE2 and SCORE2(OP) tables. A single Vascular age table may be used for all European Countries

**Lay summary:** There are no vascular age tables derived from SCORE2 and SCORE2(OP), so the aim of this study is to construct new vascular age tables derived from SCORE2 and SCORE2(OP) in a similar way to how vascular age tables derived from SCORE were published in 2010.

- Four vascular age tables have been obtained corresponding to countries with low, moderate, high and very high cardiovascular risk.
- Vascular age does not need calibration, so a universal vascular age table has been created.

## INTRODUCTION

The vascular risk tables of the European SCORE (Systemic Coronary Risk Estimation) project were published in 2003. SCORE tables calculated the risk of dying from cardiovascular causes based on age, sex, being a smoker or not, the systolic blood pressure figure and the total cholesterol level ^1^.

Since then, SCORE tables have been included in the European guidelines for cardiovascular prevention ^2^ and in the subsequent European guidelines for the management of dyslipidemia^3^.

The SCORE project intended to calculate these risk tables with European data at a time when there was a large use of tables from the American Framingham Heart Study^4^. The tables from this study could be calibrated to the European countries that wanted to use them. The SCORE project provided different tables for countries with high and low cardiovascular risk, dividing Europe into two groups of countries according to this risk by using pooled data from 16 European cohorts^1^.

Another important difference between the two studies or projects was the mathematical model underlying the risk equations to calculate the tables. While the tables from North America were based on the Cox proportional hazards model, the SCORE project was based on the Weibull regression model.

Vascular age tables derived from SCORE were published in 2010 ^5^ and from 2012 the European guidelines for cardiovascular prevention and European guidelines for the management of dyslipidemia have included the concept of vascular age until now ^6,7^.

Two reasons have been invoked to include vascular age in the guidelines. On the one hand, the cardiovascular risk tables show that at earlier ages, even in the presence several risk factors, the absolute risk may be low and thus may induce a delay in the initiation or reinforcement of cardiovascular prevention measures. On the other hand, elder individuals may present a high cardiovascular risk associated with age with a relative mild contribution of other CVRF. Therefore, complementary indicators to absolute risk such as relative risk, vascular age and lifetime risk were proposed. In addition, absolute cardiovascular risk has a mathematical component that does not allow the patient to be aware of the own real risk situation, and most important, does not inform about the excess of risk attributable to treatable risk factors besides age.

If absolute risk is translated into vascular age, the patient may have a more direct understanding of the actual life implications of the absolute risk. By this mechanism, patients may be more willing to adhere to lifestyle or pharmacological measures indicated by their physicians ^10^.

To report the vascular age of a patient with a given absolute cardiovascular risk estimated by SCORE, the age at which an individual of the same sex and country would reach the same absolute risk with controlled CVRF is calculated.

The European guidelines mention the vascular age tables developed from the SCORE ^5^ and show how vascular age can be roughly estimated from the SCORE tables (Supplemental Figure 1)^2^. To do this, the age at which the lower left box (normal or controlled systolic blood pressure and cholesterol values and non-smokers) contains the same absolute risk value as the patient is looked for in the SCORE tables.

In 2021, new cardiovascular morbidity risk tables were published with a methodology similar to that of the SCORE project, thus called SCORE2 ^8^. From different European cohorts, Cox proportional hazards regression models were calculated according to 4 different risk zones (low, moderate, high and very high) and adjusted for five-year periods, according to sex. The CVRFs considered in the cardiovascular risk tables were sex, smoking, systolic blood pressure and non-HDL cholesterol levels, as well as age (from 40 to 70 years) and risk area. In parallel, tables with other cohorts for ages 70 to 90 years are published with the same methodology, called SCORE2-OP ^9^, to be applied for patients within this age range. Both tables, SCORE2 and SCORE2-OP, have been included in the 2021 European Guidelines for Cardiovascular Prevention as the reference for absolute vascular risk estimation for different age ranges. These guidelines continue to consider the possibility of informing the patient of vascular age.

SCORE2 and SCORE2-OP tables present absolute risk information in two ways: with the numerical value corresponding to the absolute cardiovascular risk and a color (green, orange and red) corresponding to the risk level (low to moderate, high and very high). In these new tables, the cut-off points that determine the level of risk depend on age, unlike the SCORE tables in which the cut-off point was constant.

At present, there are no vascular age tables derived from SCORE2 and SCORE2-OP.

The aim of this study was to calculate the vascular age tables derived from the published cardiovascular risk tables SCORE2 and SCORE2-OP for each of the 4 risk European zones.

## METHODS

### 1. Calculation of baseline cardiovascular risk curves

The SCORE2 and SCORE2-OP tables have been used in the version grouped by the four risk areas as stated in the 2021 European Cardiovascular Prevention Guidelines^6^.

From the lower left box of each five-year period of non-smoking subjects, a mathematical model was sought that relates age and absolute risk for each sex in each of the 4 risk zones that fits the data in the tables (Supplemental Figure 2).

Different regressions (linear, quadratic, cubic, potential, logarithmic and exponential) between absolute risk and age were calculated and their adjustments assessed by calculating Pearson’s regression coefficient r and the coefficient of determination r2 to select the model that best fitted. Namely, regressions were calculated for the 8 baseline conditions (in non-smokers with systolic blood pressure (SBP) of 110 mmHg and non-HDL-cholesterol of 3.45 mmol/l) corresponding to each sex in the four risk zones.

### 2. Calculation of risk curves for each box in the risk tables

With a methodology similar to that used to calculate baseline risk, with the best-fit model, 16 regressions between absolute risk and age were calculated for each box corresponding to the crossing of the 4 categories of systolic blood pressure and 4 categories of non-HDL cholesterol, in each combination of sex (2 categories), smoking (2 categories) and risk zone (4 zones), except for the 8 basal already calculated previously, summing 16*2*2*4-8=248 different regressions for non-basal data (Supplemental Figure 3).

Pearson’s regression coefficient r and the coefficient of determination r2 were calculated for each of the 248 regressions to check the goodness of the fits.

To assess the agreement of the absolute risk between the calculated regressions and the original data in the tables, risk tables were constructed with the calculated regressions and compared with the original tables using intraclass correlation coefficients.

### 3. Conversion of calculated risk to vascular age and creation of vascular age tables

Once the 256 regression models have been calculated and their fit and agreement have been checked, the risk was calculated according to the model corresponding to each box in the SCORE2 and SCORE2-OP risk tables. This risk was used in the baseline curve corresponding to the same sex and the same risk area. By clearing the age, the vascular age corresponding to each box in the tables was calculated.

If for a box “x” that corresponds to a point of one of the 248 regressions, its risk is calculated as risk_x_=F_x_(age_x_), we must look for the age at which its baseline risk (for the same sex and risk zone with the risk factors controlled) risk_0_=G_0_(agev) is the same as the one calculated. Equalizing both risks clears up equality as:

F_x_(age_x_)= G_0_(age_v_)

Vascular age = age_v_= G ^-1^(F_x_(age_x_))

With all the vascular age data calculated for all the boxes, the vascular age tables were created. In addition, the original color of each box corresponding to the absolute cardiovascular risk category (low to moderate, high or very high cardiovascular risk) was preserved so both absolute risk tables and vascular age tables maintain the same color code for risk classification according to biological age proposed in the guidelines.

Once the vascular age tables have been calculated, the agreement between pairs of tables was calculated using the intraclass correlation coefficient.

## RESULTS

### 1. Calculation of baseline cardiovascular risk curves

The mathematical model that best adjusts baseline risk in the 4 risk zones by sex is the exponential model: Basal risk_0_=a_0_+b_0_*exp(c_0_*age). In populations at very high cardiovascular risk, a continuous model composed of two exponential models corresponding to the SCORE2 and SCORE2-OP tables was calculated, with interpolation transition between them for each sex.

The Pearson regression coefficients obtained for each sex and area varied between 0.998 and 0.9999 and the coefficients of determination between 0.997 and 0.9999 (Supplemental Table 1).

With the obtained regression equations, the baseline risk curves for each risk population and sex were calculated (Supplemental Figure 4)

### 2. Calculation of the risk curves for each box in the risk tables

248 exponential regressions of the form risk_x_=a_x_+b_x_*exp(c_x_*age_x_) for each box “x”, whose Pearson’s r coefficients varied between 0.99404 and 0.99997. Four of them were 0.994 and the rest (244) above 0.996. The coefficients of determination varied between 0.98812 and 0.99994, with only 4 out of 248 lower than 0.99.

To assess the agreement of between the regression-calculated data and the original data, new vascular risk tables were constructed for each risk area. Intraclass correlation coefficients ranged between 0.998 and 1.

### 3. Conversion of calculated risk into vascular age and creation of vascular age tables

To calculate the vascular age in each box of the tables, the absolute risk was initially calculated by means of the corresponding regression from among the 248 regressions (according to risk area, sex, age, systolic blood pressure, non-HDL cholesterol and smoking): risk_x_=a_x_+b_x_*exp(c_x_*age_x_)

With the regression corresponding to the baseline situation (Baseline risk_0_=a_0_+b_0_*exp(c_0_*age_0_) for the same risk zone and sex, the two risk expressions were equated: a_x_+b_x_*exp(c_x_*age_x_)=a_0_+b_0_*exp(c_0_*age_0_)

Vascular age was thus obtained by clearing up the equation: Vascular age= age_0_=Ln((a_x_-a_0_+b_x_*exp(c_x_*age_x_)/b_0_)/c_0_

By repeating the calculation for all boxes, vascular age tables were created for SCORE2 and SCORE2-OP for each of the four risk populations according to sex with a format similar to the SCORE2 and SCORE2-OP risk tables. The color of each box of the risk tables in the vascular age tables has been preserved to maintain the information on the stratified risk level as low, moderate and high, and each box contains the value of the vascular age calculated from the 248 regressions (Figures 1 to 4).

**Figure 1.**
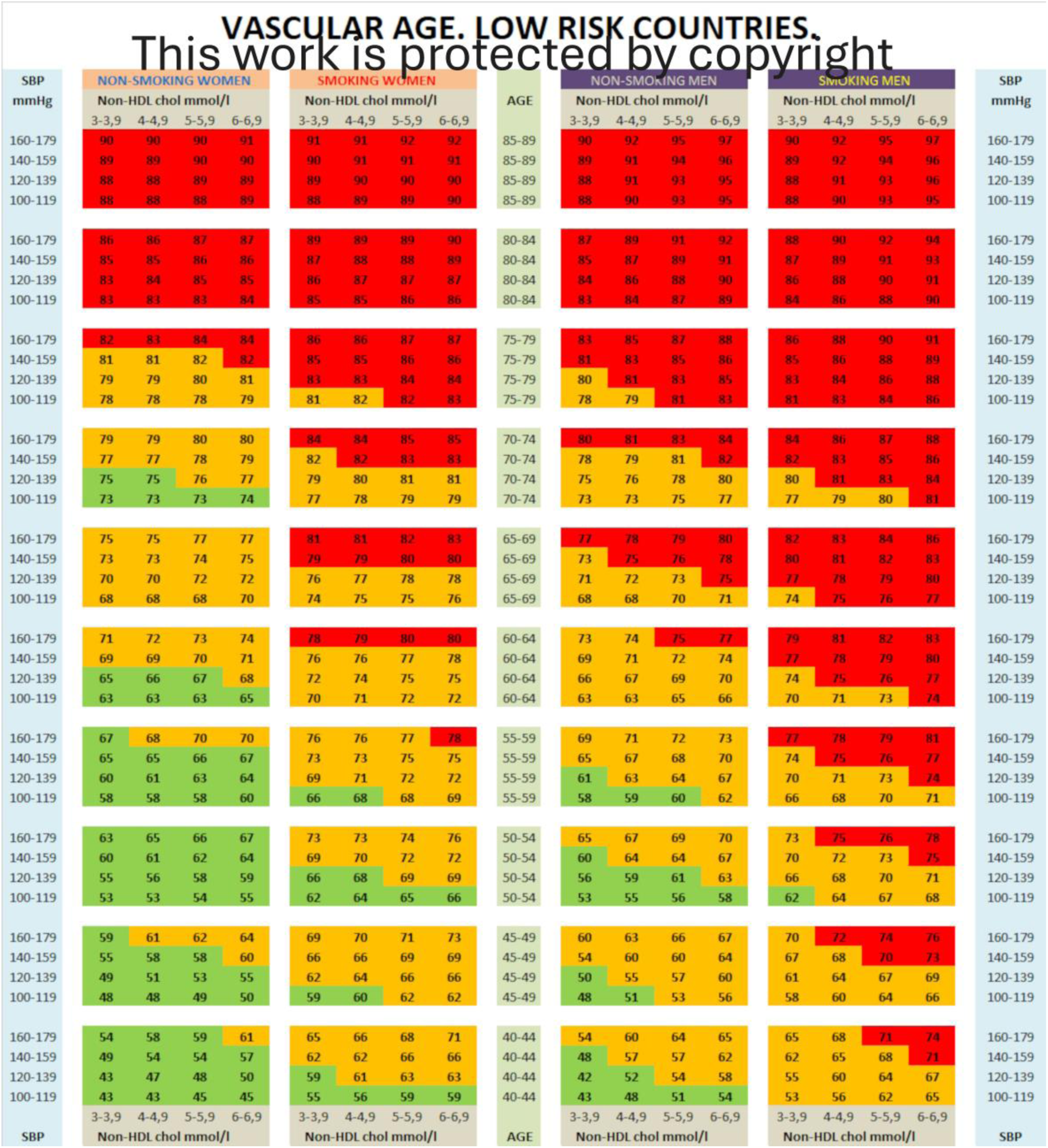
Table of vascular age for low-risk countries. Green boxes: low risk. Orange boxes: moderate risk. Red boxes: high risk.

**Figure 2.**
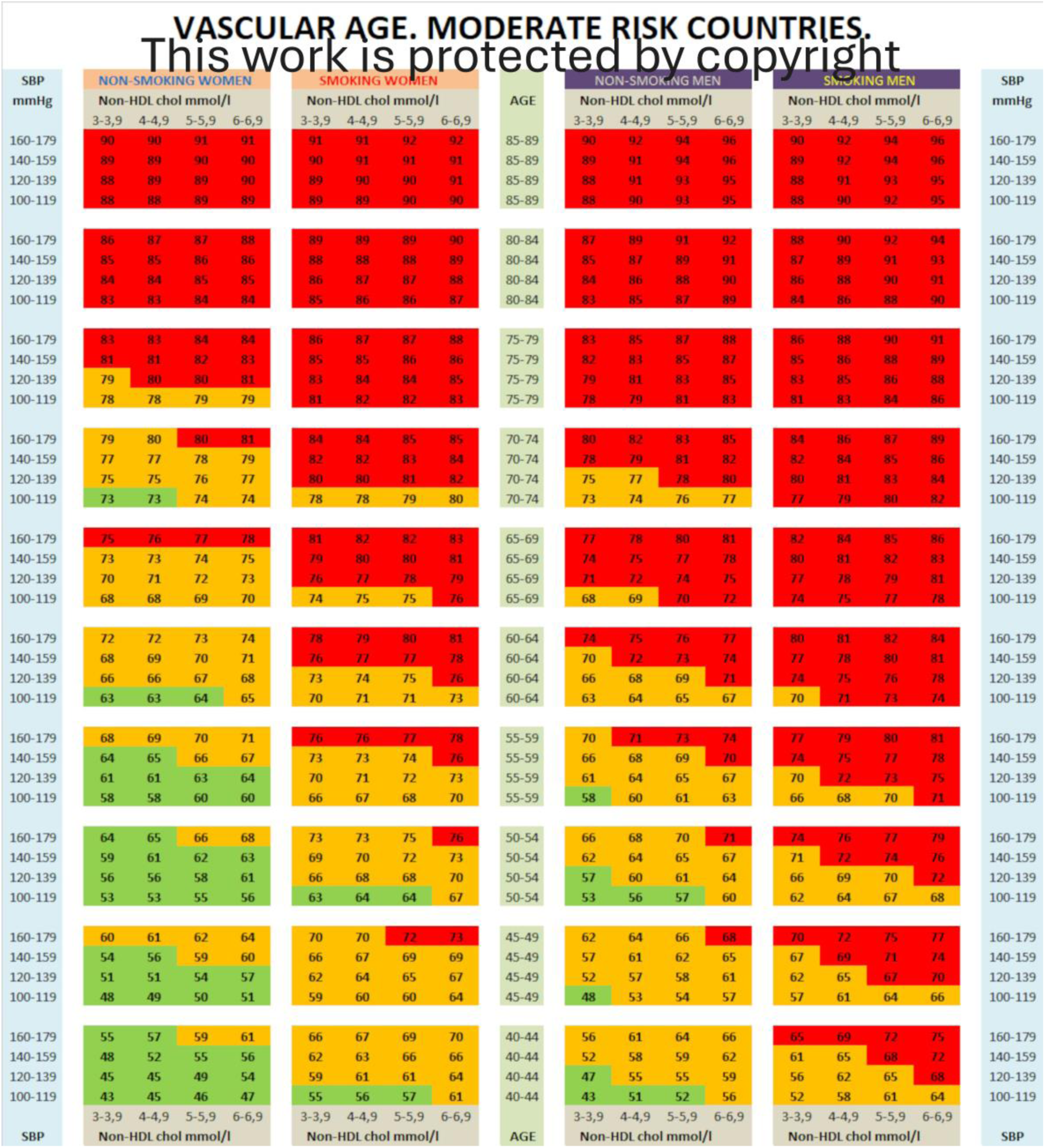
Table of vascular age for moderate-risk countries. Green boxes: low risk. Orange boxes: moderate risk. Red boxes: high risk.

**Figure 3.**
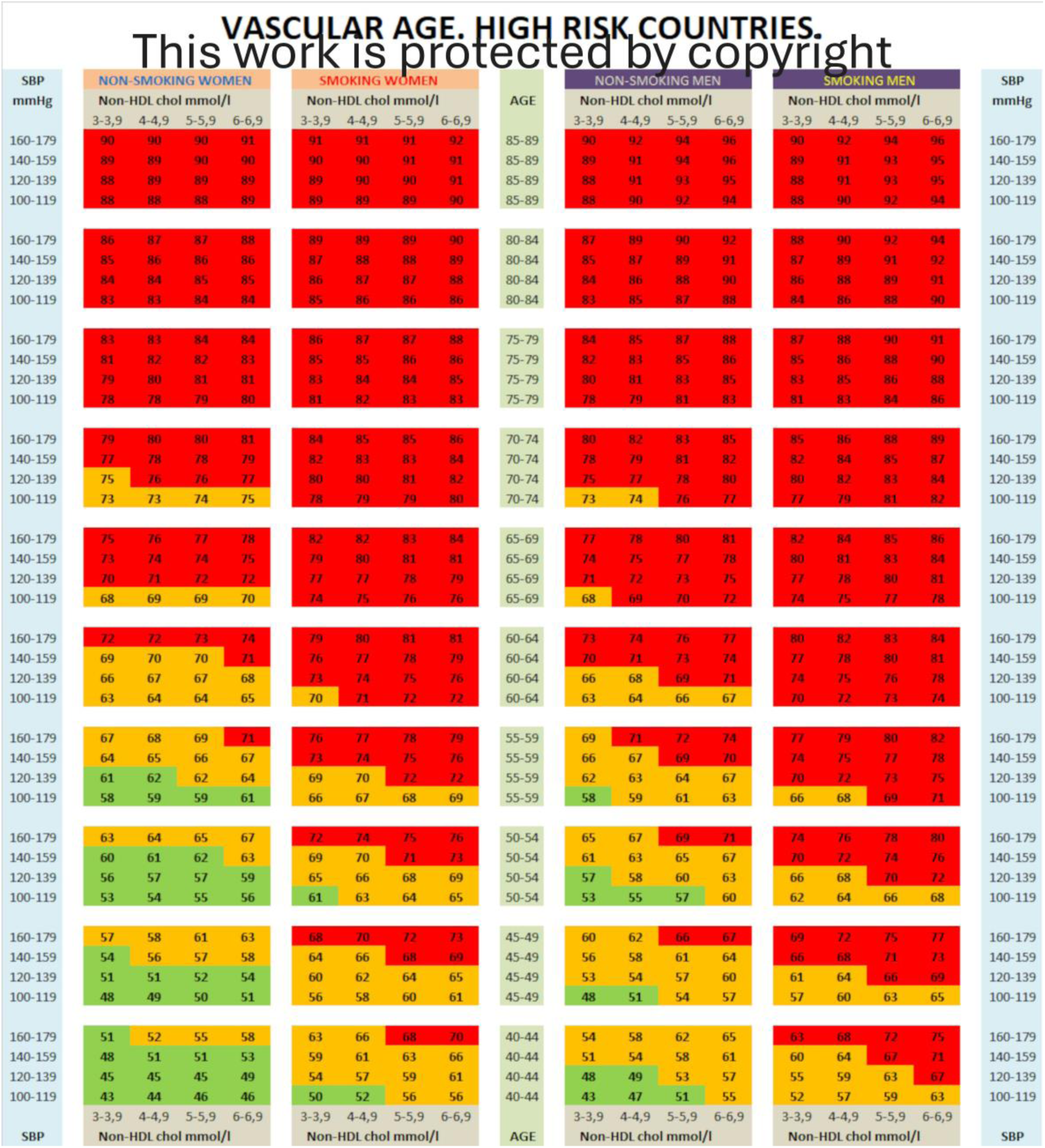
Table of vascular age for high-risk countries. Green boxes: low risk. Orange boxes: moderate risk. Red boxes: high risk.

**Figure 4.**
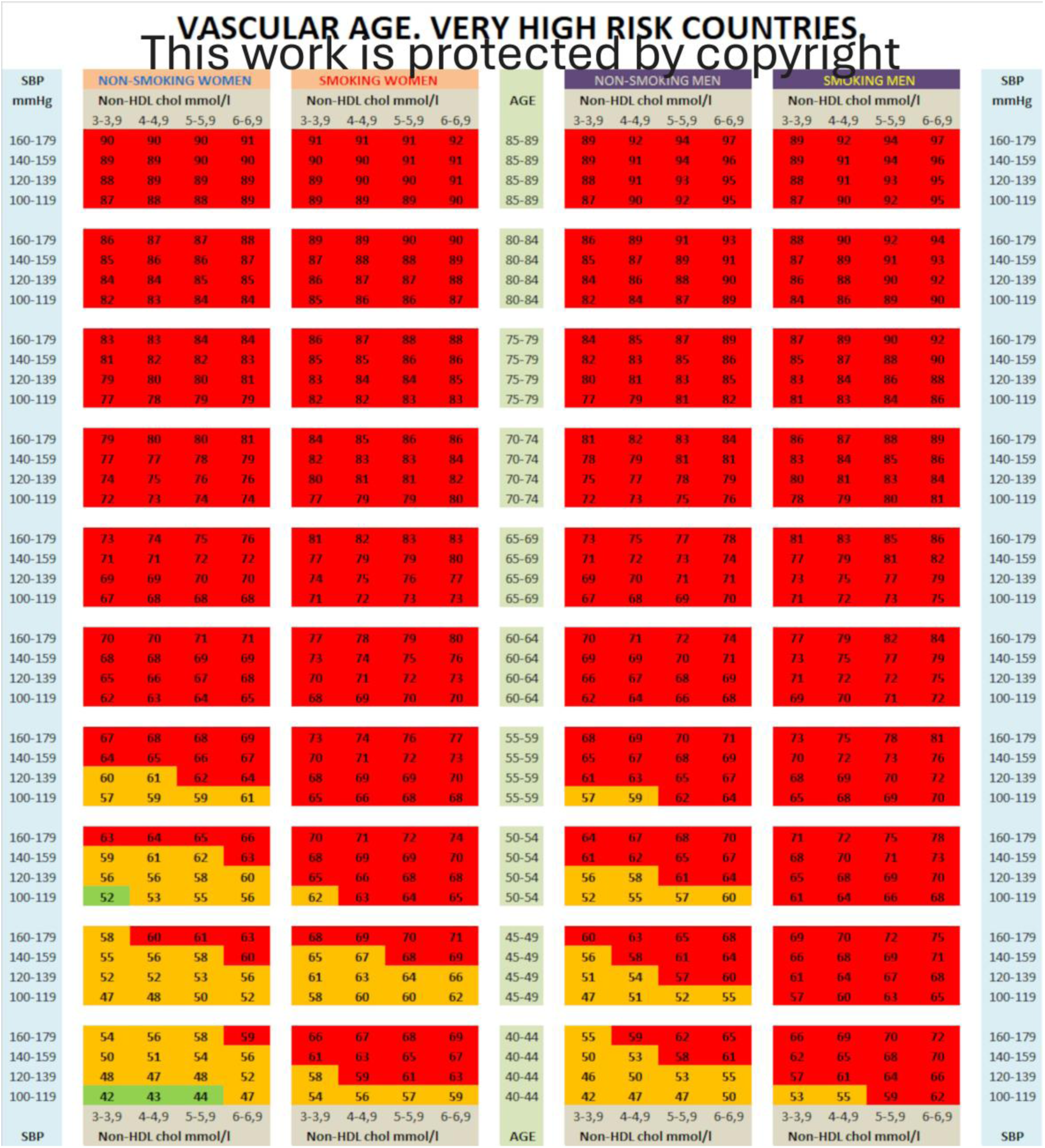
Table of vascular age for very high-risk countries. Green boxes: low risk. Orange boxes: moderate risk. Red boxes: high risk.

The agreement between the different vascular age tables according to the risk populations has intraclass correlation coefficient values of 0.992 to 0.998 (Supplemental Table 2), with the vascular age table for low-risk populations being the one that has the greatest agreement with the rest. The overall agreement of the 4 tables has been 0.993.

Given the high agreement obtained between the different vascular age tables, a new table has been calculated with the mean of the most disparate data in each box between the 4 vascular age tables. This new average vascular age table had a concordance with the specific tables for risk populations between 0.992 and 0.998 (Supplemental Table 2), so it could be used by any population (Figure 5) in the 4 European risk areas.

**Figure 5.**
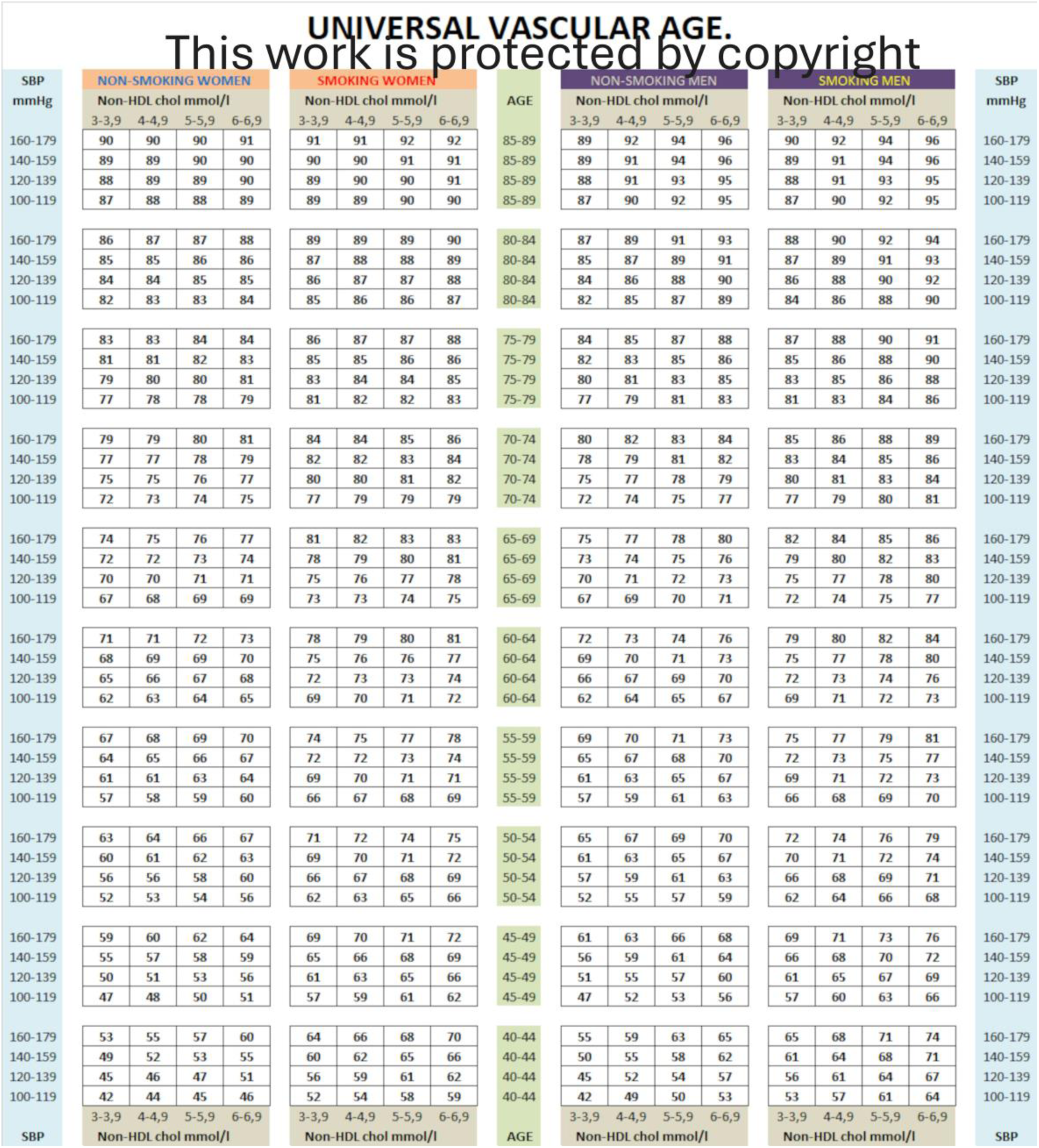
Universal table of vascular age.

## DISCUSSION

The concept of vascular age is included in the new European guidelines for cardiovascular prevention as in previous editions, but as the new cardiovascular risk tables SCORE2 and SCORE2-OP replace SCORE tables, it is necessary to update the vascular age tables to keep consistency.

The new SCORE2 and SCORE2-OP tables consider 4 at-risk populations instead of 2 as in the original SCORE. In addition, non-HDL cholesterol replaces total cholesterol and vascular morbidity replace mortality in the new tables. Finally, new vascular risk estimation extends the age range from 40 to 65 years old in the original SCORE tables to 40 to 90 years old if we assemble the SCORE2 and SCORE2-OP tables. These are additional reasons by which an update of the vascular age tables is required to keep consistency with the updated vascular risk estimation tables

The mathematical model used for the construction of the risk tables with the adjustment by age brackets of 5 years (SCORE2^8^ y SCORE2-OP^9^) improve the predictive capacity of the tables but make it impossible to create a continuous function derived directly from the original model in order to calculate vascular age. This has forced us to look for a continuous model by means of a regression with the baseline values of each table by sex and risk zone (8 baseline regressions). The regressions obtained are extraordinarily consistent with the original data of the tables with coefficients of determination greater than 0.99, i.e., more than 99% of the variability of the vascular risk calculated in the risk tables is explained or adjusted with the regressions obtained (Supplemental Table 1). These high values show that the methodology used is valid for the purpose of calculating baseline risk curves (Supplemental Figure 4) by sex and risk populations necessary to calculate the vascular age tables (Figures 1 to 4), in a similar way as was done with the curves based on SCORE. Vascular age is a projection of a person’s calculated risk at the age at which the baseline risk is reached.

Then, each risk value of each box of the SCORE2 and SCORE2-OP risk tables could have been projected onto the corresponding baseline curve to obtain the vascular age in each box. However, this methodology would have led to imprecise vascular age values: the risk values in the tables are rounded to a percentage integer value and therefore two contiguous boxes with the same absolute risk would give two identical vascular ages. This would imply a very imprecise rounding of vascular age, especially at young ages, where the use of vascular age is more interesting and where small changes in risk imply relatively larger changes in vascular age. In Supplemental Figure 4, it can be seen that at young ages the risk changes little as the years advance (the curve is very horizontal) while at older ages for vascular age to change much there must be large changes in risk (more vertical part of the curve). This circumstance has forced us to perform the calculation of 248 regressions on the data from the risk tables to obtain equations that allow a more accurate estimation of vascular age. The validity of this approximation is demonstrated by the high coefficients of determination obtained with figures greater than 0.99 in 244 regressions and 0.988 in the remaining 4. In other words, approximately 99% of the data contained in all the risk tables are adjusted to the calculated regression models. This validity is confirmed by the high agreement between the data in the original tables and those obtained with the 256 regressions (intraclass correlation coefficients greater than 0.99). Interestingly, this accuracy is higher than the actual way we calculate biological age in years, which implies an average error of estimation of about 0.5/age (i.e. higher than 5% for individuals younger than 100 years old). This inaccuracy is further increased if SCORE tables are constructed for 5-year intervals.

The conversion of absolute risk into vascular age has been possible by the use of exponential regressions in all cases. In contrast SCORE (based on transcendental equations) ^1^, does not have an algebraic resolution^5^.

Finally, once the vascular age tables for the four risk populations have been constructed, the agreement of the vascular age calculated with each of the tables was assessed. The intraclass correlation coefficients obtained in the concordance study by pairs of tables were between 0.992 and 0.998 (Supplemental Table 2), very high values with extraordinary agreement. This agreement was also seen with the vascular age tables of the SCORE with an ICC of 0.997^5^.

Therefore, vascular age calibration is not as necessary as absolute risk calibration. This absence of the need for calibration has also been demonstrated with the vascular age derived from the Framingham study^11^. In order to create a global European vascular age table, i.e. applicable to any European population, a vascular age table has been calculated with the average of the extreme values found in each equivalent box in the vascular age tables of the 4 populations. This European global vascular risk table has CCIs between 0.992 and 0.998 when compared to the 4 tables, similar to the one calculated with the SCORE. Again, this concordance is better than our standard way of measuring age in years. In addition, a global European table allows an approach for individuals whose classification in one of the risk areas is questionable (i.e. a 45 year old Romanian (very high risk) living in Spain (low risk) for the last 15 years).

The extraordinary concordance of vascular age tables from different areas of the world is not necessarily surprising, since ‘vascular age estimation’ is a way of estimating risk from classical risk factors excluding age, i.e. risk conveyed by tobacco, blood pressure and lipids. Many vascular risk calculators include this risk factors as independent coefficients in exponential regressions, thus independent of baseline vascular risk.

Interestingly, these 3 ‘classical risk factors’ may be addressed by lifestyle changes as well as pharmacological approaches. This approach may offer an estimation in terms of expected benefits in time free of cardiovascular events by subtracting actual estimation of vascular age with the estimated vascular age if risk factors are better controlled. The improved ‘vascular’ age may help to stimulate patients to engage in stop smoking initiatives or adherence to intense lipid lowering therapies (Graphical Abstract).

Our proposal has important limitations. First, all calculations have been made from reference tables where age and vascular risk factors are displayed as intervals. However, this limitation is inherent to the original tables. Taking this into account, the extraordinary concordance of individual calculated values and those estimated by our table suggests that our table provide similar accuracy as the original SCORE2 tables. In addition, a number of vascular risk modulators (intensity of smoking, obesity, presence of other clinical conditions such as inflammatory diseases, socioeconomic conditions…) are not included in the tables, as it is the case for SCORE2/SCORE2 OP tables that feed vascular age estimations.

The use of interpolation for values in the transition from SCORE2 to SCORE2-OP is an arbitrary choice. However, we believe is a reasonable and simplified practical approach for patient follow-up rather than jumping from SCORE2 to SCORE2-OP that may provide false ‘improvements’ in vascular estimation as patients reach the age of 70 years.

Our proposal allows to move from the possibility of calculating vascular age in a rough way by looking for the grid with a lower left box with the same risk, to the reality of presenting the exact value of vascular age and in the same box the vascular risk categorized as low, moderate and high with a color code. The high regression and intraclass correlation coefficients demonstrate the validity of the results, allowing the inclusion of the new vascular age tables in clinical practice guidelines. Vascular risk age tables may be an additional instrument to estimate the ‘excess’ of risk attributable to classical risk factors and help to involve patients into priorities for RF control. The global risk age estimation for all European regions may also be useful for citizens from any region or moving across Europe.

## Data Availability

All data produced in the present work are contained in the manuscript

## Supplemental Figures and Tables

**Supplemental Figure 1.**
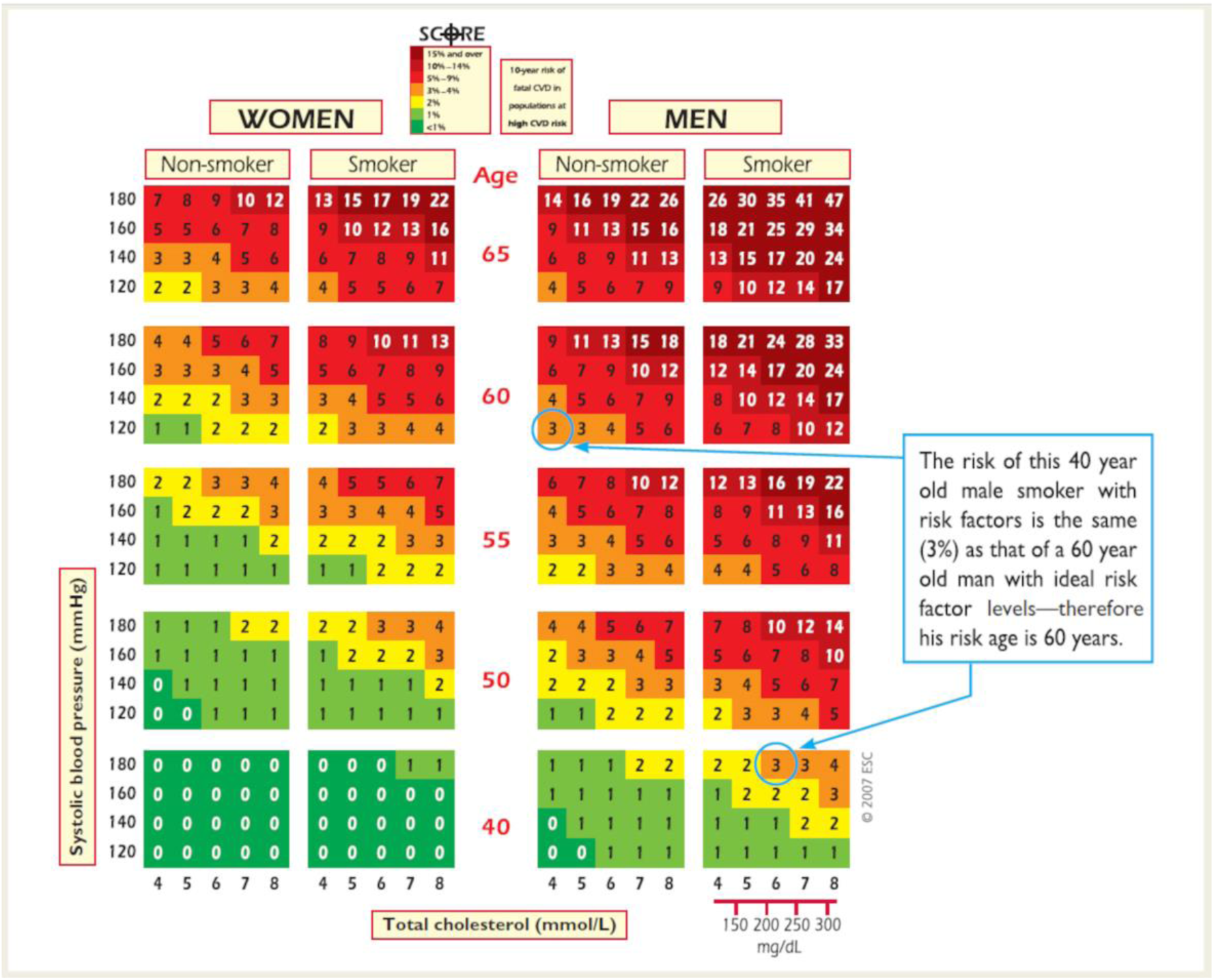
Approximate estimation of vascular age with SCORE tables^2^.

**Supplemental Figure 2.**
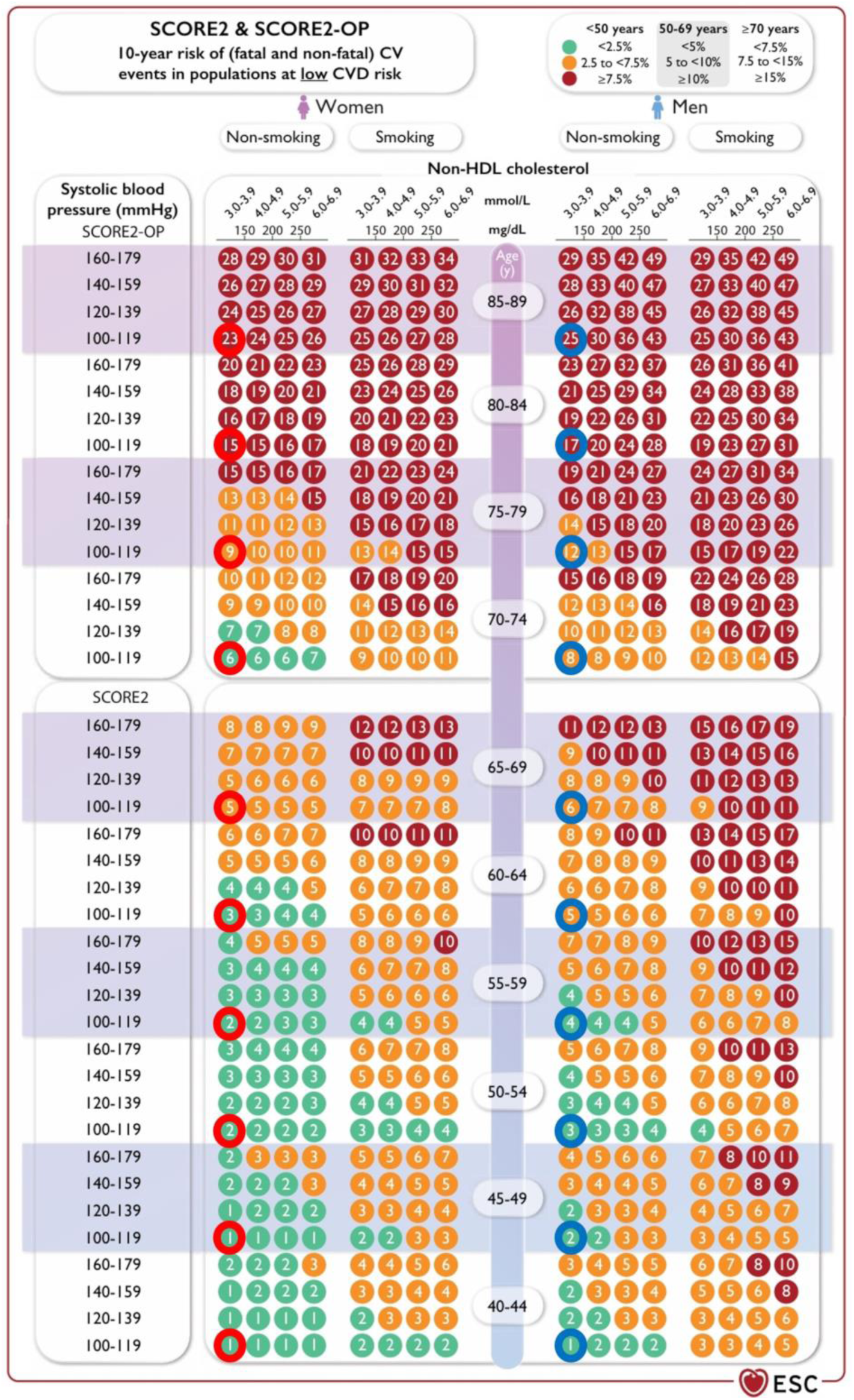
Lower-left boxes (circled) used to calculate the regression models in baseline conditions between absolute risk and age (modified from Visseren FIJ et al^6^).

**Supplemental Figure 3.**
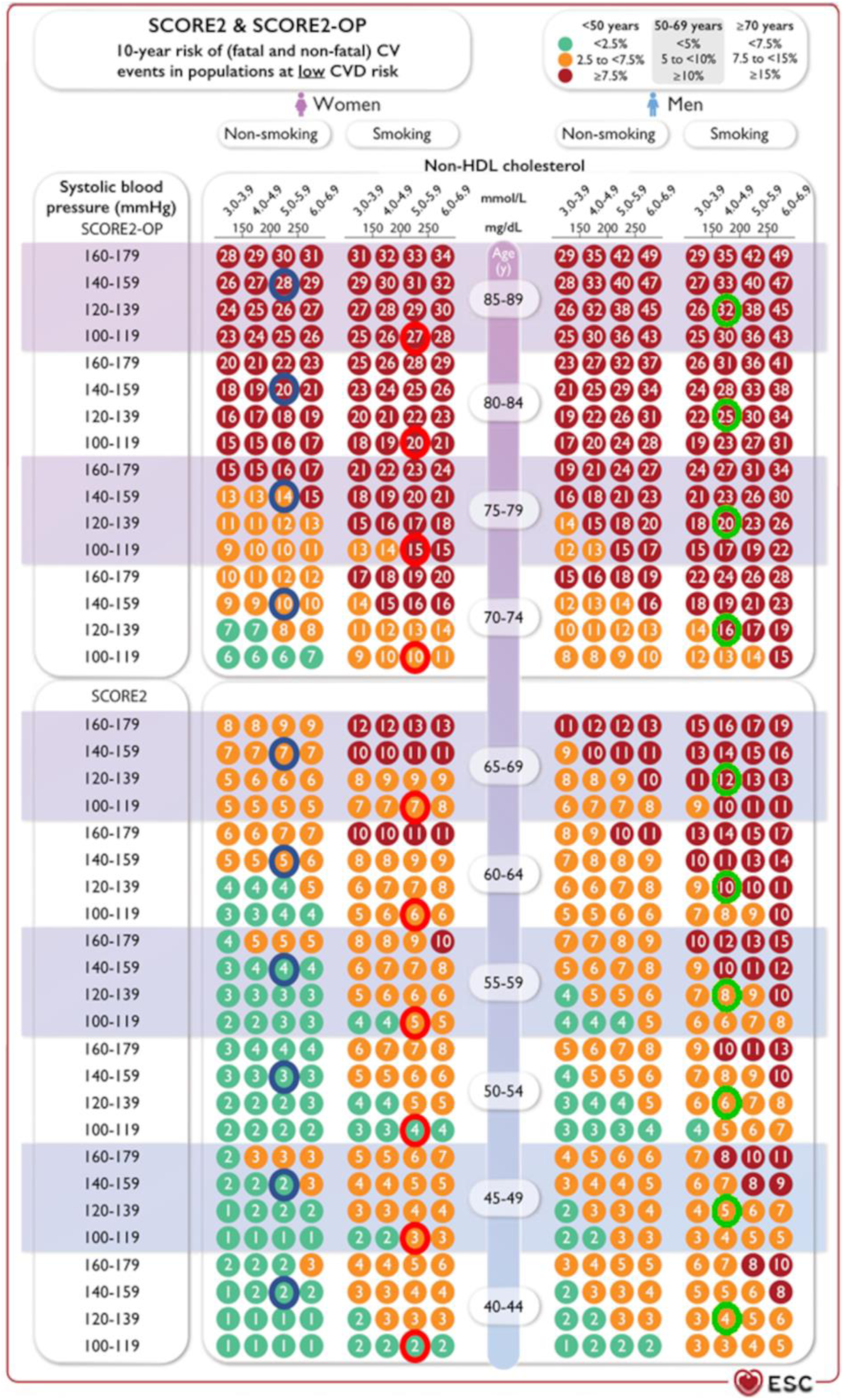
Example of boxes used to calculate 3 regressions (blue, red and green circles) of the 248 non-basal regressions between absolute risk and age (modified from Visseren FIJ et al^6^).

**Supplemental Figure 4.**
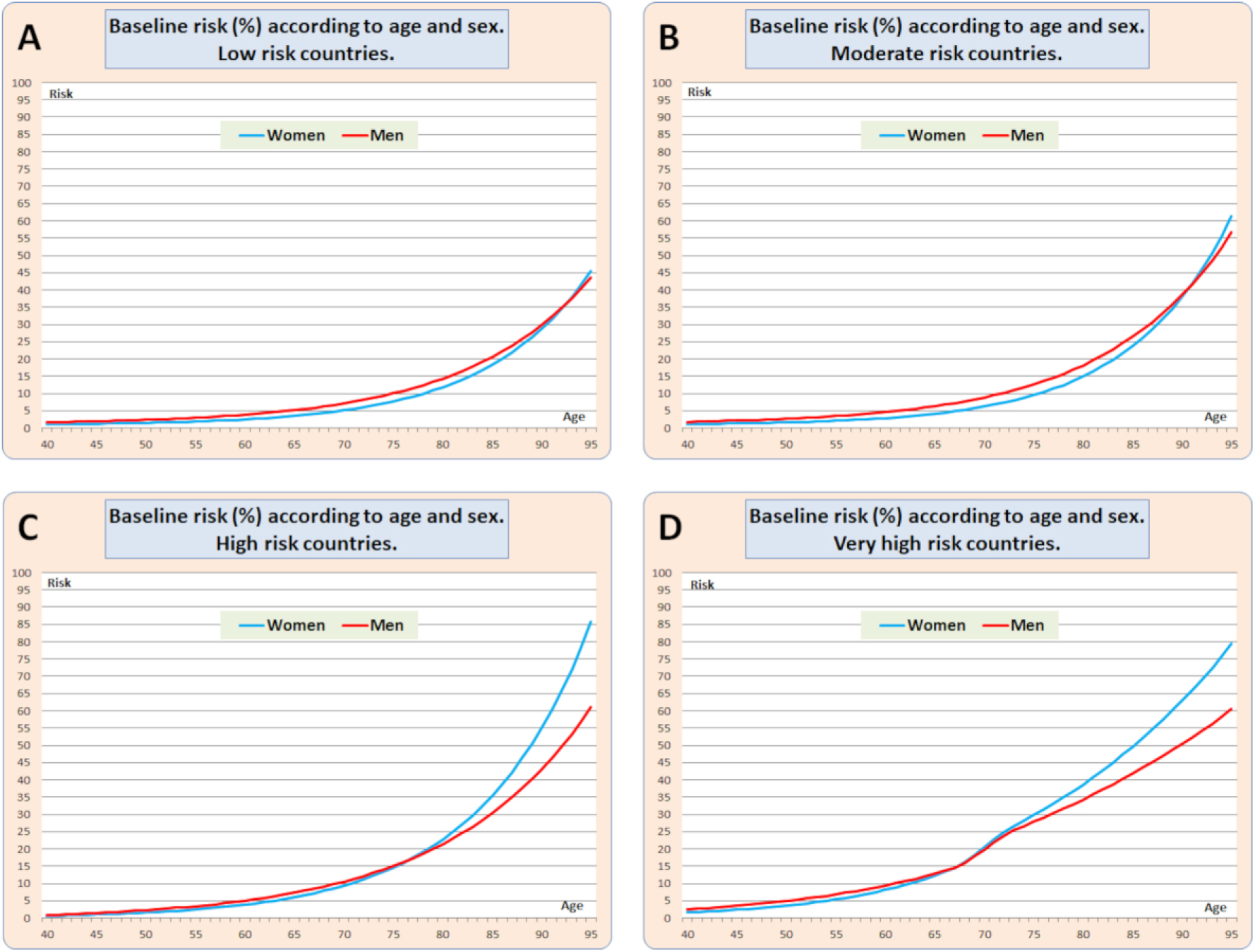
Baseline cardiovascular risk (with risk factors controlled) (SCORE2 and SCORE2-OP) according to age for low (A), moderate (B), high (C), and very high (D) cardiovascular risk countries.

**Supplemental Table 1.** Pearson’s regression coefficients and coefficients of determination of exponential regressions for baseline risk according to the model Risk = a+b*exp(c*age) for each risk population and sex.

| | | Pearson's r-coefficient | Coefficient of determination $r^2$ |
| --- | --- | --- | --- |
| Low risk | Women | 0.999 | 0.997 |
|  | Men | 0.998 | 0.997 |
| Moderate risk | Women | 0.999 | 0.998 |
|  | Men | 0.999 | 0.999 |
| High risk | Mujeres | 0.999 | 0.999 |
|  | Men | 0.999 | 0.999 |
| Very high risk | Mujeres | 0.9999 | 0.9999 |
|  | Men | 0.9999 | 0.9999 |

**Supplemental Table 2:** Concordance analysis: Intraclass correlation coefficients between each pair of vascular age tables.

|  | Moderate | High | Very high | Universal |
| --- | --- | --- | --- | --- |
| Low | 0,998 | 0,997 | 0,994 | 0,998 |
| Moderate |  | 0,997 | 0,992 | 0,997 |
| High |  |  | 0,993 | 0,997 |
| Very high |  |  |  | 0,992 |

